# Effectiveness of RMSSD-Based Adaptive Music Therapy (Skitii) in Reducing Treatment-Related Anxiety in Head and Neck Cancer Patients: Protocol for a Randomized Controlled Trial

**DOI:** 10.64898/2026.05.13.26353099

**Authors:** Prabha Adhikari, M Dinesh, Vijayalakshmi Subramaniam, Tejesh Krishna, B Ashra, Chirag Bhikamchand Jain

**Author notes:** **Corresponding Author:** Dr. Prabha Adhikari, Department of Oncology, Yenepoya Medical College Hospital, Yenepoya (Deemed to be University), Deralakatte, Mangalore, Karnataka 575018, India. **Trial Registry** CTRI/2025/11/116732, registered November 2025. **CDSCO Licence** SW/MD/MD13/2026/000003509 — Class B Medical Device. **Ethics Approval** Yenepoya Ethics Committee 1 (YEC-1), Protocol No. YEC-1/2026/006, Version 3.0. **Sponsor** Mindful Gurukul Private Limited | |. **Study Site** Dept. of Radiation Oncology, Room 3B-14, Yenepoya Medical College Hospital, Mangalore 575018. **Study Duration** August 2026 – February 2027 (8 months total). **Protocol Version** Version 2.0 (manuscript) based on clinical protocol Version 3.0, March 18, 2026.

## Abstract

**Background:** Head and neck cancer (HNC) patients experience clinically significant anxiety in 65-85% of cases during active treatment. Anxiety is associated with treatment dropout rates reaching 26% in some cohorts, with each discontinuation resulting in suboptimal outcomes and estimated losses of INR 4-5 lakh per patient. Current supportive care lacks personalized, real-time non-pharmacological interventions. Skitii is an investigational medical device (CDSCO Class B, Licence SW/MD/MD13/2026/000003509) that uses continuous RMSSD monitoring via Polar H10 chest sensor to select music in real-time, targeting parasympathetic recovery (RMSSD >=30ms).

**Methods:** Prospective, open-label, randomized controlled superiority trial (1:1 allocation) at Yenepoya Medical College Hospital, Mangalore, India. Adults aged 18-75 years with confirmed HNC (any subsite, Stage I-IV) undergoing curative-intent radiotherapy and/or chemotherapy with baseline anxiety (HADS-A >=8 and <=18) will be enrolled. Participants are randomized to Skitii adaptive music therapy (30-minute sessions, 3 times daily, 4 weeks) or static music therapy control. Skitii employs a two-phase algorithm: Phase 1 (0-2.5 minutes) uses heart rate as a stress proxy; Phase 2 (2.5-20 minutes) uses RMSSD to adapt music every 2.5 minutes when physiological state changes by >=20%, targeting RMSSD >=30ms. Primary endpoint: HADS-A score change from baseline to Week 4. Secondary endpoints: treatment dropout rate, resting RMSSD, HADS-Depression, EORTC QLQ-C30, GAD-7, PHQ-9 and device validation (ICC >=0.90 vs Kubios HRV Standard). Sample size: 70 (35 per arm), powered at 80% to detect a 2.5-point HADS difference (SD=3.8, alpha=0.05, 15% dropout). Analysis: ANCOVA, intent-to-treat.

**Discussion:** This is the first randomized controlled trial evaluating RMSSD-based adaptive music therapy in cancer patients, and the first clinical investigation of Skitii as a CDSCO-registered investigational medical device. The active control design (static music therapy) isolates the specific effect of the adaptive algorithm from music exposure alone. A mandatory device validation sub-study with 15 healthy volunteers precedes patient enrollment. If positive, results will support CDSCO Class B device certification, future CE Mark (EU MDR 2017/745), and FDA De Novo clearance applications.

**Trial Registration:** Clinical Trials Registry - India: CTRI/2025/11/116732, registered November 2025.

## 1. Background and Rationale

### 1.1 Clinical Problem

Head and neck cancers (HNC) represent approximately 30% of all cancer diagnoses in India and 6% globally, with India accounting for one-third of the worldwide burden. Treatment is aggressive, typically involving curative-intent radiotherapy with or without concurrent chemotherapy, and is associated with profound physical and psychological morbidity. Patients face fears of disfigurement, loss of speech and swallowing function, and significant treatment toxicities.

Treatment-related anxiety has profound clinical and economic consequences. Studies demonstrate that 65-85% of HNC patients experience clinically significant anxiety (HADS-A >=8) during radiotherapy and/or chemotherapy. High baseline anxiety correlates with poor treatment adherence, with dropout rates reaching 26% in some cohorts. Each premature treatment discontinuation results in suboptimal clinical outcomes and substantial economic losses, estimated at INR 4-5 lakh per patient in lost healthcare revenue and wasted resources. Despite this, psychological support remains systematically under-resourced in Indian oncology settings.

### 1.2 Limitations of Current Music Therapy Approaches

Music therapy has demonstrated effectiveness in reducing anxiety across diverse medical populations. A Cochrane systematic review by Bradt et al. (2016) analysed 52 trials involving 3,731 participants and found that music interventions significantly reduced anxiety (SMD -0.60, 95% CI - 0.83 to -0.37) and improved quality of life in cancer patients.

However, a critical limitation of all existing music therapy approaches is that they use static, pre-selected playlists without any real-time physiological feedback. Music is delivered uniformly regardless of the patient’s actual autonomic state at any moment during the session. This ignores well-documented individual and intra-session variability in stress response and misses the opportunity to match the therapeutic stimulus to the patient’s current physiological need.

### 1.3 Heart Rate Variability as an Objective Stress Biomarker

Heart rate variability (HRV), specifically the RMSSD (root mean square of successive differences of R-R intervals), is a validated, non-invasive measure of autonomic nervous system balance. RMSSD reflects parasympathetic (vagal) tone: higher values indicate greater parasympathetic activity and lower stress; values below 30ms indicate elevated sympathetic dominance and psychological stress, as established by the Task Force of the European Society of Cardiology (1996).

In cancer patients, reduced HRV correlates directly with higher anxiety levels. Chiang et al. (2013) demonstrated that advanced cancer patients show significantly reduced RMSSD, with the strongest correlation observed with reported anxiety scores (r = -0.62, p<0.001). Zhou et al. (2016) showed that HRV predicts cancer-related fatigue and survival. Despite this evidence, no published clinical trial has used real-time RMSSD monitoring to actively drive a personalized therapeutic intervention in cancer patients.

### 1.4 The Skitii System: Innovation and Regulatory Status

Skitii (Mindful Gurukul Private Limited, Mumbai) is an investigational medical device that addresses the gap between static music therapy and objective physiological monitoring. Skitii has received CDSCO Class B medical device licence (SW/MD/MD13/2026/000003509) under Medical Device Rules 2017, Rule 3(18), classified as a moderate-risk non-invasive software-based medical device for therapeutic intervention in a psychological condition.

The system uses a Polar H10 chest-worn heart rate sensor to continuously capture R-R intervals at 1000Hz, calculates RMSSD in real-time, and uses this signal to select music from a structured therapeutic library targeting parasympathetic recovery (RMSSD >=30ms). The patent-filed two-phase adaptive algorithm resolves the cold-start problem inherent in real-time HRV monitoring. Phase 1 (0-2.5 minutes) uses instantaneous heart rate as a validated predictive proxy for stress state using HR thresholds: >95 bpm (high stress, grounding music), 85-95 bpm (moderate stress, calming), 70-84 bpm (normal resting, restorative), <70 bpm (relaxed, uplifting). Phase 2 (2.5-20 minutes) transitions to RMSSD-based real-time adaptation, updating music every 2.5 minutes when RMSSD category changes by >=20%. The algorithm learns individual patient responses across sessions, excluding tracks that consistently fail to improve RMSSD.

The Polar H10 has been extensively validated against medical-grade ECG systems, achieving ICC >0.99 for R-R interval measurement (Gilgen-Ammann et al., 2019; Hernando et al., 2023). Offline endpoint analysis uses Kubios HRV Standard (free academic version), used in over 1,000 peer-reviewed publications following Task Force (1996) standards.

This trial is the first randomized controlled trial of RMSSD-based adaptive music therapy in any clinical population, and specifically the first in cancer patients. The clinical investigation is conducted under CDSCO MD-12 (Application No. MD12/2025/1247) and MD-22 (Application No. MD22/2026/0089) regulatory pathways, in compliance with ISO 14155:2020, ICH-GCP E6(R2), and ICMR National Ethical Guidelines 2017.

## 2. Objectives

### 2.1 Primary Objective

To reduce treatment-related anxiety scores by >=20% (measured by HADS-A) in HNC patients receiving Skitii adaptive music therapy compared to static music therapy control, measured as the difference in mean change from baseline to Week 4.

### 2.2 Secondary Objectives

- To achieve >=15% reduction in treatment dropout rates (premature discontinuation of radiotherapy or chemotherapy for non-medical reasons) at Week 4 and Week 8
- To validate Skitii device accuracy in measuring RMSSD with ICC >=0.90 and mean bias <2ms compared to Kubios HRV Standard (gold standard)
- To compare resting RMSSD at Week 4 between arms
- To assess depression using HADS-D, GAD-7, and PHQ-9
- To evaluate quality of life using EORTC QLQ-C30
- To measure blood cortisol as a physiological stress biomarker at baseline, Week 2, Week 4
- To assess participant satisfaction and acceptability (>=70% positive response rate on 10-item satisfaction scale)
- To document device-related adverse events

### 2.3 Hypotheses

#### Primary

H1: The Skitii adaptive music therapy group shows significantly greater reduction in HADS-A score from baseline to Week 4 compared to static music therapy (expected mean difference >=2.5 points, SD=3.8).

#### Null

H0: There is no significant difference in HADS-A score change from baseline to Week 4 between the two groups.

## 3. Methods

### 3.1 Study Design

Prospective, randomized, controlled, open-label, parallel-group, single-centre superiority trial with 1:1 allocation ratio. Blinding of participants and clinical staff is not feasible given the nature of the intervention. The biostatistician conducting primary analysis will be blinded to group allocation until database lock. The trial is registered with CTRI (CTRI/2025/11/116732) and conducted under CDSCO investigational medical device regulations (MD-12 and MD-22 pathway).

### 3.2 Study Setting and Duration

Department of Radiation Oncology, Room 3B-14, Yenepoya Medical College Hospital, Deralakatte, Mangalore, Karnataka, India (575018). Study duration per participant: 12 weeks total — 4-week intervention period (Weeks 0-4) followed by 8-week observational follow-up (Weeks 4-12). Primary endpoint assessment at Week 4. Secondary endpoints at Weeks 4, 8, and 12. Overall study timeline: August 2026 to February 2027.

Intervention room specifications: private, air-conditioned (12×10 ft), reclining chair, adjustable lighting, soundproofing, emergency call button, dedicated research room with restricted access. Sessions scheduled Monday-Friday, 9:00 AM-5:00 PM, aligned with participants’ routine hospital visits for radiotherapy or chemotherapy to minimise additional travel burden.

### 3.3 Eligibility Criteria

#### Inclusion Criteria

- Age 18-75 years; written informed consent obtained after mandatory 24-hour cooling period
- Histologically confirmed HNC (oral cavity, oropharynx, hypopharynx, larynx, nasopharynx, salivary glands), any stage (I-IV, AJCC 8th edition)
- Curative-intent treatment with radiotherapy and/or chemotherapy, minimum 3 weeks planned treatment remaining at enrollment
- ECOG Performance Status 0-2; life expectancy >3 months
- Baseline anxiety: HADS-A >=8 and <=18 (clinically significant but not severe enough to require immediate psychiatric intervention)
- Able to communicate in English, Hindi, or Kannada (validated questionnaire languages)
- Access to smartphone (Android 8.0+ or iOS 12+); willing to wear Polar H10 chest strap for 30 minutes 3 times daily

#### Exclusion Criteria

- Active cardiac arrhythmia requiring antiarrhythmic medication, recent MI (<3 months), severe heart failure (NYHA III-IV), implanted pacemaker or ICD, chronic atrial fibrillation with uncontrolled ventricular rate
- Severe cognitive impairment (MMSE <20), active psychotic disorder, severe depression with suicidal ideation (HADS-D >15), uncontrolled epilepsy, active substance use disorder
- Concurrent enrollment in another interventional clinical trial
- Palliative care intent only; planned treatment transfer within study period
- Severe bilateral hearing loss; documented allergy to medical adhesives, silicone, or polyester; active skin conditions at electrode placement site; pregnancy
- Geographic inaccessibility (>100km from hospital without reliable transportation)

### 3.4 Interventions

#### Intervention Arm: Skitii Adaptive Music Therapy

Participants receive 30-minute music therapy sessions, 3 times daily (morning, afternoon, evening), for 4 weeks (up to 84 sessions total). Sessions conducted in the dedicated research room using the Skitii mobile application with Polar H10 chest sensor. Phase 1 (0-2.5 minutes): heart rate measured in first 30 seconds, music selected from corresponding therapeutic category based on HR thresholds. Phase 2 (2.5-20 minutes): RMSSD calculated from 2.5-minute sliding window, music adapted every 2.5 minutes if RMSSD category changes by >=20%. The hysteresis threshold prevents disruptive rapid switching. Target physiological state: RMSSD >=30ms. The algorithm learns patient-specific responses across sessions, permanently excluding tracks that fail to improve RMSSD.

#### Control Arm: Static Music Therapy

Participants receive music sessions at the same frequency, duration, and setting. A pre-determined playlist of 20 instrumental tracks (10 Category 2 calming, 60-70 BPM; 10 Category 3 restorative, 70-80 BPM) plays in a fixed alternating sequence. The Polar H10 sensor is worn and HRV data is collected but not used to adapt music. Participants retain the ability to manually skip tracks they find uncomfortable (standard music therapy patient agency). This arm isolates the effect of the adaptive algorithm by controlling for music exposure duration, frequency, and setting.

### 3.5 Randomization

Computer-generated randomization with permuted blocks (sizes 4 and 6, randomly varied). Stratification by three treatment-stage strata: Stratum A (pre-treatment, target n=24), Stratum B (early treatment 1-4 weeks, target n=24), Stratum C (mid-late treatment 5+ weeks, target n=22). This stratification ensures balanced distribution of baseline anxiety levels, as anxiety typically varies by treatment stage. Allocation concealed using sequentially numbered, opaque, sealed envelopes stored in a locked cabinet, opened only after baseline assessments are complete. Randomization sequence generated by an independent biostatistician not involved in recruitment.

### 3.6 Outcomes and Assessments

#### Primary Outcome

Change in HADS-Anxiety subscale score (range 0-21; >=8 indicates probable anxiety) from baseline to Week 4. Administered by trained research coordinator in participant’s preferred language using standardized script.

#### Secondary Outcomes

- Treatment dropout rate: premature discontinuation of planned radiotherapy or chemotherapy for non-medical reasons at Weeks 4 and 8
- Resting RMSSD (ms) at Week 4 — 5-minute supine recording processed by Kubios HRV Standard
- HADS-Depression subscale, GAD-7, PHQ-9 at Weeks 4 and 8
- EORTC QLQ-C30 global health status and functional scales at Weeks 4 and 8
- Blood cortisol (serum, morning 8-10am) at baseline, Week 2, Week 4
- Device validation: ICC between Skitii real-time RMSSD and Kubios offline RMSSD across 70 participants (target ICC >=0.90, mean bias <2ms)
- Participant satisfaction: 10-item Likert scale at Week 4 (intervention group only)
- Device-related adverse events throughout study

#### Assessment Schedule

| Instrument | Baseline | Week 2 | Weeks 4, 8, 12 |
| --- | --- | --- | --- |
| HADS-A (primary) | Yes | Yes | Yes |
| HADS-D | Yes | — | Yes (Wks 4, 8) |
| GAD-7 / PHQ-9 | Yes | — | Yes (Wks 4, 8) |
| EORTC QLQ-C30 | Yes | — | Yes (Wks 4, 8) |
| Resting RMSSD | Yes | Yes | Yes (all) |
| Satisfaction scale | — | — | Week 4 (intervention) |
| Adverse events | — | Continuous | Continuous |

### 3.7 Device Validation Sub-Study

Prior to main trial enrollment (July 2026, Weeks 1-2), a mandatory validation sub-study will be conducted with 15 healthy volunteers. Each volunteer undergoes a single 5-minute session with simultaneous Skitii (real-time RMSSD) and Kubios HRV Standard (offline gold-standard) recording. Success criteria (all must be met): Pearson r >0.95, ICC >0.90, mean bias <2ms, 95% limits of agreement within +/-5ms, no significant difference on paired t-test (p>0.05). If criteria are not met, the algorithm will be recalibrated and validation repeated before any cancer patient is enrolled. A formal validation report will be approved by the PI before first enrollment.

### 3.8 Sample Size

Sample size calculated for primary endpoint HADS-A change baseline to Week 4. Expected mean reduction: intervention -4.2 points, control -1.7 points (difference 2.5 points); pooled SD 3.8 points; 80% power; two-sided alpha 0.05. Required per group: 30. Adjusted for 15% dropout: 35 per group. Total: 70 participants. Feasibility confirmed: Yenepoya Medical College Hospital treats 50-60 new HNC patients monthly; estimated 70% eligibility and 60% consent rate makes recruitment over 4-5 months achievable.

### 3.9 Statistical Analysis

#### Primary Analysis

ANCOVA for HADS-A change from baseline to Week 4. Model includes: baseline HADS-A score (covariate), treatment group (fixed effect), treatment stage stratum (fixed effect), Group x Stratum interaction term. Intent-to-treat population. Missing data handled by multiple imputation (m=50, predictive mean matching). Results reported as adjusted mean difference, 95% CI, and Cohen’s d effect size. Two-sided alpha=0.05. Sensitivity analysis: per-protocol population; Mann-Whitney U if normality violated.

#### Secondary Analyses

Treatment dropout: Chi-square test. Device validation: ICC (two-way mixed, absolute agreement), Bland-Altman plots, Pearson correlation. Quality of life: repeated measures ANCOVA. Cortisol: repeated measures ANCOVA. All tests two-sided, alpha=0.05. Longitudinal trajectory: mixed-effects models with treatment x time interaction. Within-stratum sensitivity analyses for primary outcome.

#### Interim Analysis

Independent Data Safety Monitoring Board (DSMB) reviews at 25% enrollment (n=18) and 50% enrollment (n=35). Safety stopping rule: device-related SAE rate >10%. Futility stopping rule: conditional power <20% for primary endpoint.

### 3.10 Safety Monitoring

Three-tier monitoring structure: Tier 1 (Research Assistant) — present in room throughout each session, monitors device and participant comfort; Tier 2 (Trained Nurse) — daily review of session logs, 24-hour post-session check-in calls; Tier 3 (Principal Investigator, Dr. Prabha Adhikari) — weekly data review, mandatory face-to-face assessment at Weeks 2 and 4. Remote monitoring by Mindful Gurukul technical team via automated backend alerts for sensor disconnection, algorithm errors, or data transmission failures.

Expected harms and management: chest strap skin irritation (5-10%, managed with hypoallergenic alternative strap); headphone discomfort (2-3%, managed with volume reduction or speaker option); music-triggered distressing memories (3-5%, session stopped immediately, study psychologist contacted); worsening depression with suicidal ideation (<2%, immediate psychiatric referral, study discontinuation). All adverse events classified per ICH-GCP E6(R2) criteria. Serious adverse events reported to YEC-1 within 24 hours and CDSCO within 7 calendar days per Medical Device Rules 2017.

### 3.11 Ethical Considerations

Ethics approved from Yenepoya Ethics Committee 1 (YEC-1), Protocol No. YEC-1/2026/006, Version 3.0, March 18, 2026. Conducted per Declaration of Helsinki, ICMR National Ethical Guidelines 2017, ICH-GCP E6(R2), ISO 14155:2020, and Medical Device Rules 2017. All participants provide written informed consent after a mandatory 24-hour cooling period — implemented to prevent rushed decisions, enable family consultation, and reduce therapeutic misconception in this potentially vulnerable population. Comprehension assessed via 5 teach-back questions before consent signing. Participants explicitly informed that study participation does not affect their cancer treatment.

Privacy and dignity protocols established for electrode placement, including mandatory female nurse and chaperone for female participants, verbal consent before each placement, and cultural sensitivity accommodations. Independent patient advocate (Hospital Social Worker) available throughout the trial. Ethics Committee contact provided to all participants for confidential complaint reporting.

### 3.12 Data Management

Two-database architecture: Database 1 (identifiable information — name, phone, MR number) accessible by Research Coordinator only, hosted on MongoDB; Database 2 (research data — Study ID, outcomes, HRV data, session logs) accessible by PI and analysts, hosted on AWS Mumbai region (ap-south-1) with automated backup to Singapore region. Encrypted master key table (AES-256) maps Study IDs to MR numbers, requires dual-password access from both PI and Hospital IT administrator.

Study ID format: SKIT-YMC-[Stratum]-[Sequential]-[Random] (e.g., SKIT-YMC-A-023-X7K). All network transmission uses TLS 1.3. 100% source data verification for primary endpoints; 20% random verification for all other data. Data retained per CDSCO requirements: identifiable data 5 years, research data 10 years, signed consent forms 15 years per ICH-GCP E6(R2).

## 4. Discussion

### 4.1 Significance

This trial addresses a critical gap in oncology supportive care: the absence of physiologically responsive, real-time non-pharmacological interventions for cancer-related anxiety. Skitii is the first medical device to close this gap by using continuous HRV monitoring to drive music selection, creating a closed-loop therapeutic system that responds to each patient’s autonomic state in real-time.

The significance extends across clinical, regulatory, and commercial dimensions. For patients, a device that responds physiologically represents a qualitatively different experience from passive music playback. For oncology departments, Skitii offers a scalable supportive care tool that does not require clinical psychology staffing. For regulators, this trial provides the clinical evidence base for CDSCO Class B device certification, and will support future CE Mark (EU MDR 2017/745) and FDA De Novo clearance applications.

### 4.2 Strengths

- First RCT of RMSSD-based adaptive music therapy in any clinical population
- First clinical trial of Skitii as a CDSCO-registered investigational medical device
- Active control arm (static music therapy) isolates the specific effect of the adaptive algorithm
- Mandatory device validation sub-study before patient enrollment
- Three-stratum randomization controls for treatment-stage-related baseline anxiety variation
- DSMB provides independent safety oversight at 25% and 50% enrollment
- Mandatory 24-hour cooling period and comprehensive dignity protocols protect vulnerable participants
- Objective physiological primary endpoint (RMSSD) alongside validated patient-reported outcome (HADS-A)
- Cost-effective technology: Polar H10 (validated consumer device) and Kubios HRV Standard (free academic software)
- Pre-registered and pre-published protocol prevents outcome reporting bias

### 4.3 Limitations

- Open-label design: blinding of participants not possible given the adaptive nature of the intervention; patient expectation effects cannot be fully controlled
- Single-centre study: results may not generalise across different oncology centres, patient populations, or cultural music preferences
- 4-week primary endpoint: longer-term effects beyond the treatment period will require follow-up studies
- RMSSD measurement depends on adequate Polar H10 signal quality; patients with excessive movement artefact may require session exclusion
- Music library reflects specific cultural and therapeutic choices; individual music preferences vary

### 4.4 Future Directions

If positive, this trial will provide the evidence base for a multi-centre confirmatory RCT across Indian oncology centres, CDSCO Class B device certification, CE Mark application under EU MDR 2017/745, and FDA De Novo clearance. Secondary analyses will examine HRV trajectory patterns, individual music preference-response relationships, and treatment adherence correlations. The Skitii algorithm architecture is designed for expansion to other cancer types, treatment settings, and clinical applications where autonomic regulation and stress management are relevant.

## 5. Trial Status

This manuscript represents Version 2.0, based on clinical protocol Version 3.0 (March 18, 2026). The trial is in the pre-study phase: CDSCO MD-13 licence received (SW/MD/MD13/2026/000003509), CTRI registration confirmed (CTRI/2025/11/116732), YEC-1 ethics resubmission Version 3.0 approved on March 31st, 2026 (Protocol No. YEC-1/2026/006) with site permission. Device validation sub-study planned for July 2026. First patient enrollment anticipated August 2026. Study completion and data analysis expected January-February 2027.

## Data Availability

This is a study protocol. No dataset has been generated or analysed yet. De-identified data from the completed trial will be made available upon reasonable request to the corresponding author 6 months after publication of the primary results, subject to a Data Use Agreement and Yenepoya Ethics Committee approval. The CTRI registration (CTRI/2025/11/116732) is publicly accessible at ctri.nic.in.

## 6. Declarations

### Ethics Approval and Consent

Ethics approval received from the Institutional Ethics Committee 1 (YEC-1) of Yenepoya Medical College Hospital, Yenepoya (Deemed to be University), Deralakatte, Mangalore, Karnataka, India (Protocol No. YEC-1/2026/006, Version 3.0, March 31, 2026). Conducted per Declaration of Helsinki and ICMR National Ethical Guidelines 2017. Written informed consent will be obtained from all participants after a mandatory 24-hour cooling period prior to enrolment.

### Consent for Publication

Not applicable. No individual patient data is included in this protocol manuscript.

### Availability of Data

This is a study protocol. No dataset has been generated or analysed yet. De-identified data from the completed trial will be made available upon reasonable request to the corresponding author 6 months after publication of the primary results, subject to a Data Use Agreement and YEC-1 approval. CTRI registration (CTRI/2025/11/116732) is publicly accessible at ctri.nic.in

### Authors’ Contributions

PA (Principal Investigator): study conception, protocol design, clinical oversight, ethics submissions, manuscript drafting. DM (Co-Investigator): clinical protocol review, patient recruitment oversight. VS (Co-Investigator, Vijayalakshmi Subramaniam): psychological assessment oversight, questionnaire administration coordination. TK (Co-Investigator, Oncology): oncology protocol review, patient safety. AB (Research Coordinator): protocol operationalisation, data management, questionnaire administration. CJ (Sponsor Representative): device specification, algorithm design, regulatory strategy, manuscript review. All authors reviewed and approved the final manuscript.

## Acknowledgements

The authors thank the Department of Radiation Oncology at Yenepoya Medical College Hospital for institutional support and provision of the dedicated research room (Room 3B-14). We acknowledge the Yenepoya Ethics Committee 1 (YEC-1) for their rigorous review process that strengthened the protocol through three iterations. AI-assisted tools were used in the initial drafting of sections of this manuscript under full human oversight and revision; the authors take complete responsibility for all content as required by ICMR guidelines.

## Appendix: SPIRIT 2013 Checklist Note

A completed SPIRIT 2013 checklist will be provided as a supplementary file at the time of peer-reviewed journal submission. All SPIRIT items are addressed within this protocol. The trial is registered with CTRI (CTRI/2025/11/116732, registered November 2025) as required by ICMJE guidelines.

